## Supplementary material for "National snakebite project on capacity building of health system on prevention and management of snakebite envenoming including its complications in selected districts of Maharashtra and Odisha in India: a study protocol": S1 Ethics Approval and Funding Document

**ICMR - NATIONAL INSTITUTE FOR RESEARCH IN REPRODUCTIVE HEALTH**
Jehangir Merwanji Street, Parel, Mumbai-400 012
**NIRRH ETHICS COMMITTEE FOR CLINICAL STUDIES**


Recognized by: Strategic Initiative for Developing Capacity in Ethical Review (SIDCER), Forum for Ethics Review Committees in Asia and the Western Pacific Region (FERCAP) for its compliance with international and local standards in ethical review

**CHAIRPERSON**
Dr. Padmavathy Menon

**MEMBERS**

Dr. Y. S. Nandanwar
Dr. Sandeep Bavdekar
Dr. Sivakami Muthusamy
Prof. Shubhada Chiplunkar
Dr. Ketki Kulkarni
Dr. Shirish Joshi
Dr. Vikrant Bhor
Dr. Bhakti Pathak
Adv. Ajay Shinde
Mrs. Minakshi Bansode

**ALTERNATE MEMBER SECRETARY**
Dr. Vikrant Bhor

**MEMBER SECRETARY**
Dr. Beena Joshi

Ref.: D/ICEC/Sci-194/209/2021 14th November 2021

Dr. Rahul Gajbhiye
Dept. of Clinical Research
ICMR-NIRRH

Sub.: Reference to the project 'ICMR National snakebite project (INSP) on capacity building of health system on prevention and management of snakebite envenomation including its complications', Version 1.2 dated 01/09/2021

Project No.:393/2020 PI: Dr. Rahul Gajbhiye

Dear Dr. Gajbhiye

The Ethics Committee acknowledges and approves the final copy of the amendment version 1.2 dated 01/09/2021 along with participant information sheet and informed consent documents (English) which was received on 9th December 2021.

Please note that any changes to the proposal / Participant Information Sheet / informed consent form should have prior approval by the ethics committee before being implemented.

Due date for submission of completion report is after 12/08/2022

Sincerely,

Dr. Beena Joshi
Member Secretary

Dr. Padmavathy S. Menon
Chairperson

आई.सी.एम.आर - क्षेत्रीय आयुर्विज्ञान अनुसंधान केन्द्र
ICMR - REGIONAL MEDICAL RESEARCH CENTRE, BHUBANESWAR
INSTITUTIONAL HUMAN ETHICS COMMITTEE
IRB No: ECR/911/Inst/OR/2017
Federalwide Assurance # FWA00027286

***Protection of Human Subjects***
***IEC Certification/ Declaration of Exemption***

Chairman
Prof. Dr. Jadunath Prasad Das
Clinician

Member
Prof. Aruna Kumari Misra
Basic Scientist, Life Science
Prof. Dr. Prasanna Kumar Dash
Medical Scientist
Mrs. Kasturika Patnaik
Socially Aware Person
Dr. Pramod Kumar Acharya
Clinician
Dr Sisir Kumar Mahapatra
Clinician
Sri Santanu Das
Legal Expert
Prof. Rita Ray
Social Scientist
Prof. Dr. Sudhanshu Sekhar Mishra
Basic Medical Scientist
Dr. Anna S. Kerketta
Basic Medical Scientist

Member Secretary
Dr. Prakash Kumar Sahoo
Basic Scientist

**Ref No: ICMR-RMRCB/IHEC-2021/79** **Date: 18/08/2021**

1. **Request Type**
   - ☐ Original
   - ☐ Continuation
   - ☐ Exemption
2. **Type of Programme**
   - ☐ Research ☐ Contract Research
   - ☐ Institutional Activity - Out Patient Care/ Surveillance/ Referral & Outbreak Investigation
   - ☐ Collaborative ☐ Fellowship ☐ Others
3. **Proposal Identification No.**
   - **Institutional ID:**
   - **Funding Agency & ID (If any):** *ICMR*
4. **Title of Research or Activity**: " *ICMR National Snakebite Project (INSP) on capacity building of health system on prevention and management of snakebite envenomation including its complications".*
5. **Study site:** *Khurda block in Khurda district & Kashipur block in Rayagada district.*
6. **Principal Investigator:** *Dr. Amarendra Mohapatra, Scientist - F*
7. **Key Personnel at RMRC Bhubaneswar & Role:** *Dr. Sanghamitra Pati, Sci-G & Director*
8. **Certification of IRB review**
   **This activity/ proposal has been reviewed by sending emails to all members and approved by the IHEC in accordance with the Indian Council of Medical Research (ICMR), guidelines and other GCP recommendations, by full IRB review or expedited review by email on dated *03/09/2021.***
9. **Remarks (If any):**
   - **Confidentially to be maintained**
   - **Consent to be obtained**
10. **This activity/ proposal has been scrutinized and declared for exemption for review by IEC as there is less than minimal risk and no linked identifiers.**

**Dr. Prakash Kumar Sahoo**
Scientist - C
Member Secretary

Name & Designation of the Official

Signature: [signature]

Date: 03/09/2021

Dr. P. K. Sahoo, Sci-C
Member Secretary
IHEC, ICMR-RMRC-B

भारतीय आयुर्विज्ञान अनुसंधान परिषद
स्वास्थ्य अनुसंधान विभाग, स्वास्थ्य एवं परिवार कल्याण मंत्रालय, भारत सरकार

icmr
INDIAN COUNCIL OF MEDICAL RESEARCH
Serving the nation since 1911

Indian Council of Medical Research
Department of Health Research, Ministry of Health and Family Welfare, Government of India

No. 58/6/NTF-Snakebite/2019-NCD-II (pt-I) Dated: 09.9.2020

To,

The Director,
National Institute for Research in Reproductive Health (ICMR),
Jehangir Merwanji Street, Parel,
Mumbai-400012.

Sub: NTF project entitled "ICMR National Snakebite Project (INSP) on capacity building of health system on prevention and management of snakebite envenomation including its complications" under Dr. Rahul Gajbhiye, NIRRH, Mumbai.

Dear Sir,

The Director General of the Council sanctions the above mentioned research scheme initially for a period of one year from **01.10.2020** subject to extension upto the total duration specified in para 3(3) below.

The Director General of the Council also sanctions the budget allotment of **Rs. 51,03,900/- (Rupees fifty one lakh three thousand nine hundred only)** as detailed in the attached statement for the period ending the **30.09.2021.**

The grant-in-aid will be given subject to the following conditions:-

1. The payment of the grant will be made in lump-sum to the head of the Institution. The first installment of the grant will be paid generally as soon as a report regarding the commencement of the project and appointment of the staff is received by the Council. The demand for payment of the subsequent installment of the grant should be placed with the Council in the prescribed proforma attached.

2. The staff appointed on the project should be paid as indicated in the budget statement attached.

3. The approved duration of the scheme is **2 years.**

4. A report on the progress made will be submitted to the Council as and when called for.

5. The institute will maintain a separate account of the receipts and the expenditure incurred on the scheme and will furnish a utilization certificate and an audited statement of account pertaining to the grant.

**P.T.O.**

6. The other terms and conditions are indicated in Annexure-1

7. The Host Institute shall utilize the grant after following the provisions laid down in the GFRs 2017 and T.A. Rules.

8. The PI may be advised to keep the fund in a separate Saving Bank Account opened for research funds received from ICMR so as to ensure that interest earned thereon is also credited in to the Fund Account.

The receipt of this letter may please be acknowledged.

Yours faithfully,

(Ishwar Likhar)
Admn. Officer
For Director General

This issues with the concurrence of Finance Section vide **NCD/NTF/1/2020-21 dated 17.07.2020**

1. Copy together with a copy of the budget statement forwarded for information to Dr. Rahul Gajbhiye, Scientist 'D', ICMR-National Institute for Research in Reproductive Health, Jehangir Merwanji Street, Parel, Mumbai-400012.
2. Copy together with a copy of the budget statement forwarded to the **Accounts Section V** for information and necessary action.
3. Admn. Officer, (NCD) ICMR
4. Mr. Hemant (Sr. T.O.) ICMR
5. IRIS No. 2020-6008N

**For Director-General**

National Task Force Project

**Budget Statement for 1st year**
**(From 01.10.2020 to 30.09.2021)**

Sub: NTF project entitled "ICMR National Snakebite Project (INSP) on capacity building of health system on prevention and management of snakebite envenomation including its complications" under Dr. Rahul Gajbhiye, NIRRH, Mumbai.

**Budget for ICMR-NIIRH, Mumbai**

| Staff | 1st Year |
|---|---|
| 1 Scientist C (Medical) @ 64,000+NPA+HRA= Rs. 72325/- PM<br>For 2nd year @ 72,325+2220= 74,545 PM<br>PB 15600+GP 6600=22200x10% =2220 (every year) | 867900 |
| 2 Project Technical Officer @Rs. 32000 pm<br>PB 9300+GP 4600=13900x10% = 1390 (every year) | 768000 |
| **Total (A)** | **1635900** |
| **(B) Recurring (Contingencies)** | |
| IEC Training Mannual | 300000 |
| Software Development and Database maintenance | 300000 |
| **Total (B)** | **600000** |
| **(C) Travel Meeting and training** | |
| Training of Masters Trainers, TA, DA and honorarium of experts, and travel for monitoring of study sites | 500000 |
| Travel of Technical advisory group meeting expenses | 600000 |
| **Travel Total** | **1100000** |
| **Total (A+B+C)** | **3335900** |

**Budget for RMRC Bhubaneshwar, Odisha**

| **(A) Staff** | **1st Year** |
|---|---|
| 2 Project Technical Officer @Rs. 32000 pm PB 9300+GP 4600=13900x10% = 1390 (every year) | 768000 |
| **Total (A)** | **768000** |
| **(B) Contingencies (Recurring)** | |
| IEC Training Manual | 300000 |
| **Total (B)** | **300000** |
| **(C) Travel Meeting and training** | |
| Training of Masters Trainers, TA, DA and honarium of experts, and travel for monitoring of study sites | 500000 |
| Travel | 200000 |
| **Total (C)** | **700000** |
| **Total (A+B+C)** | **1768000** |
| **Grand Total of NIIRH Mumbai & RMRC Odisha** | **5103900** |

**(Rupees fifty one lakh three thousand nine hundred only)**

File No. 58/6/NTF-Snakebite/2019-NCD-II (pt-I)
Specialty: Snake Bite
Date of Start: 01.10.2020
Duration: 2 years

भारतीय आयुर्विज्ञान अनुसंधान परिषद
स्वास्थ्य अनुसंधान विभाग, स्वास्थ्य एवं परिवार कल्याण मंत्रालय, भारत सरकार

INDIAN COUNCIL OF MEDICAL RESEARCH
Serving the nation since 1911

Indian Council of Medical Research
Department of Health Research, Ministry of Health and Family Welfare, Government of India

No. 58/6/NTF-Snakebite/2019-NCD-II (pt-I) Dated: 09.09.2020

Sub: NTF project entitled "ICMR National Snakebite Project (INSP) on capacity building of health system on prevention and management of snakebite envenomation including its complications" under Dr. Rahul Gajbhiye, NIRRH, Mumbai.

----------

Reference this office letter No. 58/6/NTF-Snakebite/2019-NCD-II (pt-I) dated 17.02.2020

The Director-General, ICMR sanctions the payment of **Rs. 51,03,900/- (Rupees fifty one lakh three thousand nine hundred only)** as the full & final installment of 1st year of the grant for incurring expenditure in connection with the above mentioned research scheme. The amount of **Rs. 51,03,900/-** may be debited in the provision of **Rs. 51,03,900/-** made for the above mentioned research scheme for the current financial year.

A formal bill for **Rs. 51,03,900/-** is sent herewith for payment by RTGS to Project Account, DMRC.

(Ishwar Likhar)
Admn. Officer
For Director General

**Accounts Section, ICMR (Mandate form enclosed)**
**RFC No. NCD/NTF/1/2020-21 dated 17.07.2020**

Copy to:-
1. The Director, National Institute for Research in Reproductive Health (ICMR), Jehangir Merwanji Street, Parel, Mumbai-400012. An amount of **Rs. 51,03,900/-** of full & final installment will be sent to you in due course. The grant has been sanctioned on the conditions laid down in our letter referred to above.
2. Dr. Rahul Gajbhiye, Scientist 'D', ICMR-National Institute for Research in Reproductive Health, Jehangir Merwanji Street, Parel, Mumbai-400012
3. Admn. Officer, (NCD) ICMR
4. Mr. Hemant (Sr. T.O.) ICMR
5. IRIS No. 2020-6008N

**For Director General**
