## Supplementary material for "National snakebite project on capacity building of health system on prevention and management of snakebite envenoming including its complications in selected districts of Maharashtra and Odisha in India: a study protocol": S2 Permission for using India Map in Fig 1

Gmail **hrishikesh munshi <>**

### Request permission to publish India map (https://www.amcharts.com/svg-maps/?map=india2019) under CC BY 4.0 license

**Martynas Majeris (amCharts Support)** <> Wed, Feb 2, 2022 at 12:14 PM
Reply-To: amCharts Support <>
Cc: "" <>, hrishikesh munshi <>

##- Please type your reply above this line -##

You are registered as a CC on this help desk request (amcharts.zendesk.com/hc/requests/64089).
Reply to this email to add a comment to the request.

**Martynas Majeris** (amCharts Help & Support)
Feb 2, 2022, 8:44 GMT+2

Hi Krishna,

Thank you for reaching out.

You're welcome to include our map into your research publication.

Yours sincerely,

Martynas Majeris
amCharts

---

amCharts 5 is now available!

It's as versatile and powerful, yet much, much faster than anything else we've built before. More info: https://www.amcharts.com/javascript-charts/

**Krishna Chaaithanya**
Feb 2, 2022, 7:15 GMT+2

Dear Sir,

Seasonal greetings from India

I am a scientist working in the Indian Council of Medical Research – National Institute for Research in Reproductive and Child Health (ICMR-NIRRCH) Mumbai India. We wish to use the India map available on your website https://www.amcharts.com/svg-maps/?map=india2019 for our research publication in BMC Public Health journal. The journal requires permission from the original copyright holder to use the same.
In view of this, we would like to seek your permission in writing to use the map.
Anticipate a positive reply.

Thanking you
Krishna.

With Best Regards,

Krishna Chaaithanya Itta, Ph.D.
Scientist - 'C'
Department of Molecular Immunology & Microbiology
ICMR- National Institute for Research in Reproductive and Child Health (ICMR-NIRRCH)
Jehangir Merwanji Street,
Parel, Mumbai - 400 012.
Landline: 02224192017.
https://scholar.google.com/citations?user=wJDUX8kAAAAJ

---

This email is a service from amCharts Help & Support. Delivered by Zendesk.

[YDXY02-O820]
