## Supplementary material for "National snakebite project on capacity building of health system on prevention and management of snakebite envenoming including its complications in selected districts of Maharashtra and Odisha in India: a study protocol": S4 Participant Information Sheet

**Title of Project:** ICMR National Snakebite Project (INSP) on capacity building of health system on prevention and management of snakebite envenomation including its complications

Project Co-ordinator: Dr Smita Mahale
Former Director & Scientist G
Emeritus Scientist
ICMR-National Institute for Research in Reproductive and Child Health
J M Street, Parel, Mumbai 400012


Principal Investigator (PI): Dr Rahul Gajbhiye
Scientist D & Wellcome Trust India Alliance
Clinical & Public Health Intermediate Fellow
ICMR-National Institute for Research in Reproductive and Child Health
J M Street, Parel, Mumbai 400012


Co-Principal Investigator: Dr Himmatrao Bawaskar
Bawaskar Hospital and Research Centre,
Mahad District Raigad, Maharashtra


Co-Investigators: Dr Arun Yadav
Assistant Director of Health Services,
Directorate of Health Services, Maharashtra State
Arogyabhavan, St. Georges Hospital Campus, Fort,
Mumbai 400001


Dr Manohar Vitthalrao Bansode
Medical Superintendent
Sub District Hospital, Shahapur, District Thane
Maharashtra State, 421601


Dr Shashikant Shambharkar
District Health Officer
Gadchiroli, Maharashtra


Dr Kanna Madavi
Medical Superintendant

Sub District Hospital, Aheri
District Gadchiroli, Maharashtra


Dr Krishna Chaaithanya Itta
Scientist C
Molecular Immunology and Microbiology
ICMR-National Institute for Research in Reproductive and Child Health
J M Street, Parel, Mumbai 400012


Dr. Ranjan Kumar Prusty
Scientist B
Department of Biostatistics
ICMR-National Institute for Research in Reproductive and Child Health
J M Street, Parel, Mumbai 400012


Site Principal Investigator: Dr Amarendra Mahapatra
Scientist F
ICMR- Regional Medical Research Centre Bhubaneswar (RMRCB), Odisha, India


Co-Principal Investigator: Dr Subrata Palo
Scientist D
ICMR- RMRCBB, Odisha, India


You are invited to take part in this research study. Research is different than routine care. Routine care is based upon the best-known treatment and is provided with the main goal of helping the individual patient. The main goal of research studies is to gain knowledge that may help future patients.

This Participant Information Sheet gives you important information about the study. It describes the purpose of the study, and the risks and possible benefits of participating in the study.

Please take the time to review this information carefully. You are requested to ask for an explanation of any words you do not understand. After you have read the Participant Information Sheet, you are free to talk to the doctors/researchers about the study and ask them any questions you have. You will be given a copy of the participant information sheet and discuss it with your friends, family, or other doctors about your participation in this study.

If you have decided to take part in the study, you will be asked to sign the informed consent form which is along with this Participant Information Sheet. Before you sign the informed consent form, be sure you understand what the study is about, including the risks and possible benefits to you. You will be given a copy of the Participant Information Sheet and signed informed consent form for your future reference.

Please remember that your participation in this study is entirely voluntary. You are free to withdraw from the study at any point of time without affecting your medical care and services. Also, by signing the Consent form you have not waived off any rights as a participant.

You may please note that being in a research study does not take the place of routine physical examination or visits to your own doctor and should not be relied on to diagnose or treat any other medical problems.

**1. What is this research study about?**

Snakebite is a major public health problem in tribal and rural areas in India. Snakebite affects mainly poor, agricultural and migrant workers, tribals, hunters and mainly earning family member. Lack of awareness, inadequate knowledge of prevention of snakebite and first aid amongst community as well as peripheral health care workers, delay in receiving lifesaving treatment [anti-snake venom (ASV)] and non-availability of trained medical officers for management of snakebite contribute to higher number of deaths in Indian including Maharashtra and Odisha States.

The research study is about improving the knowledge of the community on prevention, appropriate first aid, and early transfer of snakebite patients to nearest health facility for reducing the mortality and morbidity due to snakebites in selected districts in Maharashtra and Odisha. Interviews of peripheral health care workers (ASHA, ANM, MPW) and Medical Officers will be conducted to understand the knowledge gaps. This would be crucial for planning appropriate and effective training in order to build the capacity of the health system in different zones of India. The prospective data will be collected from health facilities in all study sites for one-year period and implementation of National Snakebite Treatment Protocol (2017) will be monitored regularly.

**2. What information is known about this type of research study?**

Globally, around 81,000 to 138,000 deaths were estimated resulting from 1.8 million to 2.7 million cases of snakebite envenoming and leading to around 400 000 people with permanent physical or psychological disabilities. However, the snakebite burden is likely to be an underestimate as snakebite is not a notifiable disease and many bites and deaths go unrecorded. According to National mortality survey (2001-2003), India constitutes almost half of the global snakebite burden with estimated mortality ranging from 45,000 to 50,000 every year. Higher annual mortality rates per 100,000 population due to snakebite were reported in

13 states: Andhra Prades (6.2), Madhya Pradesh (5.9), Odisha (5.6), Jhrakhand (4.9), Bihar (4.9), Tamilnadu (4.7), Uttar Pradesh (4.6),Chhatisgarh (4.4), Karnataka (4.2), West Bengal (3.5), Gujarat (3.5), Rajasthan (3.3), Maharashtra (3.0). Jharkhand and Orissa states had higher number of deaths at ages 5–14 years whereas deaths at older ages were prominent in Andhra Pradesh, Bihar, Madhya Pradesh, and Uttar Pradesh. Female deaths were more than male deaths due to snakebites in Bihar, Madhya Pradesh, Maharashtra and Uttar Pradesh. The Director General of Health Services, Ministry of Health & Family Welfare, Government of India released National Snakebite Management Protocol in 2009. Subsequently, Ministry of Health & family Welfare (MoHFW), Government of India through National Health System Resource Centre (NHSRC) has developed Standard Treatment Guidelines, (STG, 2017).

**3. Why is this research study being done?**

Most of the people are bitten in rural and/ tribal areas during working in agricultural fields, fetching drinking water, sleeping on floor, going to school, or walking to an outdoor toilet without foot wares. Community education is therefore crucial, because many snakebites are not reported or people seek care too late. Most of these are deaths are preventable. The community is not aware about the occupational risks and simple, cost effective preventive measures can prevent a bite. There is a need to increase community awareness on prevention of snakebites, bring behavioral changes regarding occupational risk and its reduction and empower the community on first aid skills and early transfer of snakebite patients to nearest health facility to avoid delay in receiving lifesaving treatment (ASV) to reduce deaths and disabilities due to snakebite envenomation. The community is also required to be educated on several do's and don'ts regarding first aid for snakebites as recommended by National snakebite management protocol (2009) as well as STG, 2017.

In India, especially in primary health centers, the medical doctors are replaced every 6–12 months either due various reasons and reported to have poor knowledge about, and experience in, management of snake bites. Similarly, multiple protocols are followed at secondary and tertiary level health facilities mainly from western textbooks that are not appropriate for our Indian settings. There is no formal training of medical students and/or interns on snakebite management in India. There is no formal training program for Medical Officers working in public health system in India. Therefore, the present study will empower the community and public healthcare system in selected districts of Maharashtra and Odisha states on prevention and management of snakebites.

**4. How will the research study be done?**

Two years' retrospective data on snakebite will be collected. Focus group discussions will be conducted separately for males and females in each study area. The culturally appropriate IECs, training manuals for outreach health worker (in regional languages), Training manuals for Medical Officers will be developed under the guidance of National snakebite experts. The facility check assessment will also be carried out to understand ASVs distribution and utilization in selected regions of India. Interviews of outreach health care workers (ASHA, ANM, MPW) and Medical Officers will be conducted to understand the knowledge gaps. This would be crucial for planning appropriate and effective training in order to build the capacity of the health system in different zones of India. The prospective data will be collected from health facilities in all study sites for one-year period and implementation of National Snakebite Treatment protocol (2017) will be monitored regularly. The impact evaluation of the capacity building will also be carried out.

**5. Who can take part in this research study?**

Community members the in selected study blocks of Maharashtra and Odisha states above 18 years of age can participate in this study through focus group discussions. Medical Officers at health facilities can participate through interviews. All the public health facilities in the block except sub centers will be assessed.

**6. How many participants will be included for this research study?**

A total of 6 FGDs (3 males, 3 females) will be conducted in each block. Each FGD will be restricted to 8 to 16 participants. Overall, 24 FGDs will be conducted but the number of FGDs will vary according to the saturation of responses. Each block will have approximately 40 MOs; so nearly 160 MOs will be trained during the study. Training will also be provided to outreach healthcare workers. Approximately 150 workers per block will be trained so a total of 600 workers will be trained in four blocks. Actual numbers will vary and the same MOs or healthcare workers may not be available throughout the study duration as there are frequent transfers in the health system.

**7. What do you have to do if you agree to take part in the research study?**

You will have to participate in an FGD for about 60 minutes or an interview for about 45 minutes.

**8. What are the possible benefits to you by being in the research study?**

The study will not benefit you directly. However, the information provided by you regarding knowledge gaps will be useful for planning appropriate and effective training in order to build the capacity of health system in different zones of India. The experience of this research will be critical for adopting National Protocol with variation if required suitable for different geographic locations in the country. The research study will empower the health system for better management of snakebites in public health care system in selected districts of Maharashtra and Odisha states.

9. **What are the tests that will be performed on the participant/ biological sample?**

No tests will be performed during the course of the study.

**10. How long will you be in the research study?**
You will have to take part in the FGD or interview only once in the study.

**11. How long the biological samples will be stored and how will it be disposed?**
Not applicable

**12. Under what conditions will your Participation in the study be terminated?**
Your participation will be terminated only if during the course of the discussion, you decide not to continue in the study.

**13. What are the possible risks and inconveniences that you may face by being in the research study?**
There is no risk for you by participating in the FGD. The FGD is to assess knowledge gaps on awareness of venomous and non-venomous snakebites, prevention, first aid and management of snakebites.

**14. What happens if you are injured since you took part in this research study?**
Not applicable

**15. What are the other treatment options/alternatives to participation?**
Not applicable

**16.What will happen if you change your mind about participation in this research study?**
Participation in the study is voluntary, if you do not want to continue you can withdraw from participation at any time.

**17. How will your privacy and confidentiality be maintained?**
A central database to capture all the data generated during data collection phases will be developed and maintained at ICMR-NIRRCH, Mumbai. The database will have provision for real-time data entry. Qualitative data including audio recordings and transcripts will be kept separately and all the records that identify the participants will be kept confidential. Data access will be controlled independently and data will be secured behind firewalls as per the ICMR National Guidelines for Biomedical and Health Research Involving Human Participants [31]. Only designated individuals with login credentials will be able to access data. In case of data collected using pen and paper method, data entry and verification will be done independently. Data will be backed up regularly in hard drives with sufficient memory space. Only RG will have ultimate authority over trial data.

**18. Will you have to bear any Expenses or Costs by participating in the research study?**
You will not have to bear any cost for participating in the research study. There will be no incentives for participating in the study.

**19. Whom do you call if you have questions or problems?**

a. Research related
b. Regarding rights as a Participant

Please contact the researchers listed below to:
Obtain more information about the study
Ask a question about the study procedures or treatments

Dr Rahul Gajbhiye
Scientist D & Wellcome Trust India Alliance
Clinical & Public Health Intermediate Fellow
ICMR-National Institute for Research in
Reproductive and Child Health, Mumbai 400012

Or
Dr Manohar Vitthalrao Bansode
Medical Superintendent
Sub District Hosipital, Shahapur, District Thane
Maharashtra State, 421601

Or
Dr Kanna Madavi
Medical Superintendant
Sub District Hospital, Aheri
District Gadchiroli, Maharashtra

Or
Dr Amarendra Mahapatra
Scientist F
ICMR – Regional Medical Research Center
Chandrashekharpur, Bhubaneswar – 751023

Odisha, India

Or
Dr Subrata Palo
Scientist D
ICMR – Regional Medical Research Center
Chandrashekharpur, Bhubaneswar – 751023
Odisha, India


If you have questions or concerns about your rights as a research participant or a concern about the study, please feel free to address the Ethics Committee through the Ethics Office. (Please feel free to address the Ethics Committee through the Ethics Office and identify yourself by the ‘participant identification number’ as filled in your participant enrollment form – for sensitive study like HIV)

Dr. Beena Joshi
NIRRCH Ethics Committee for Clinical Studies
National Institute for Research in Reproductive and Child Health
J.M.Street, Parel. Mumbai 400 012
Tel.No.: Direct- 022-24192043/022-24192147, Board no.- 022-24192000

Time to contact- Monday- Friday (9.00am to 5.00 pm)

The Institutional Ethics Committee for Clinical Research comprises of a group of people like doctors, researchers, and community people (non-scientific) who work towards safeguarding the rights of the study participants like you who take part in research studies undertaken at the institute - National Institute for Research in Reproductive Health
Do not sign this consent form unless you have had a chance to ask questions and have received satisfactory answers to all of your questions.
If you agree to participate in this study, you will receive a signed and dated copy of this consent form for your records
