## Supplementary material for "National snakebite project on capacity building of health system on prevention and management of snakebite envenoming including its complications in selected districts of Maharashtra and Odisha in India: a study protocol": S5 Consent Form

**ICMR- NATIONAL INSTITUTE FOR RESEARCH IN REPRODUCTIVE AND CHILD HEALTH**

J. M. Street, Parel, Mumbai 400 012

### Informed Consent Form

I ___________________________________ have read / have had read the participant information sheet version no. 1.2 dated 01.09.2021 bearing page numbers 1-8 of the research study entitled **"*ICMR National Snakebite Project (INSP) on capacity building of health system on prevention and management of snakebite envenomation including its complications*"**.

The information contained in the participant information sheet regarding the nature and purpose of the study, safety, and its potential risks / benefits and expected duration of the study, and other relevant details of the study including my role as a study participant have been explained to me in the language that I understand. I have had the opportunity to ask queries, which have been clarified to my satisfaction.

I understand that my participation is voluntary and that I have the right to withdraw from the study at any time without giving any reasons for the same. This will not affect my further medical care or any legal rights.

I understand that the information collected about me during the research study will be kept confidential. The representatives of sponsor, government regulatory authority's / ethics committees may wish to examine my medical records / study related information at the study site to verify the information collected. By signing this document, I give permission to these individuals for having access to my records.

I hereby give my consent willingly to participate in this research study. I am informed that I will not be given any compensation/ reimbursement for participation in the study.

For limited readers or non-readers (Illiterate participants): I have witnessed the consent procedure of the study participant and the individual has had the opportunity to ask questions. I confirm that the individual has given consent freely.

| | |
|---|---|
| Signature of an impartial witness<br>with date<br>(Only if the participant is illiterate) | Signature/Thumb impression of<br>study participant with date |
| Name of the Witness | Name of the study participant |
| Signature of PI with date | Signature of person administering<br>the consent with date |
| Name of the PI | Name of the person administering<br>the consent |
