## Supplementary material for "National snakebite project on capacity building of health system on prevention and management of snakebite envenoming including its complications in selected districts of Maharashtra and Odisha in India: a study protocol": S6 WHO Trial Registration Data Set

**World Health Organization Trial Registration Data Set**

| **Data Category** | **Information** |
|---|---|
| Primary registry and trial identifying number | Clinical Trials Registry India no. CTRI/2021/11/038137 |
| Date of registration in primary registry | 18 November 2021 |
| Secondary identifying numbers | NA |
| Source(s) of monetary or material support | Indian Council of Medical Research<br>V. Ramalingaswami Bhawan, P.O. Box No. 4911<br>Ansari Nagar, New Delhi - 110029, India |
| Primary sponsor | Indian Council of Medical Research<br>V. Ramalingaswami Bhawan, P.O. Box No. 4911<br>Ansari Nagar, New Delhi - 110029, India |
| Secondary sponsor(s) | NA |
| Contact for public queries | Dr. Rahul K. Gajbhiye<br>Scientist D & DBT Wellcome India Alliance<br>Clinical and Public Health Intermediate Fellow,<br>ICMR-National Institute for Research in Reproductive and Child Health, Mumbai, Maharashtra, 400012 India,<br>Telephone +91 22 24192036<br> |
| Contact for scientific queries | Dr. Rahul K. Gajbhiye<br>Scientist D & DBT Wellcome India Alliance<br>Clinical and Public Health Intermediate Fellow,<br>ICMR-National Institute for Research in Reproductive and Child Health, Mumbai, Maharashtra, 400012 India,<br>Telephone +91 22 24192036<br> |
| Public title | Improving community awareness and capacity of healthcare providers for prevention and management of snakebite envenoming |

| **Data Category** | **Information** |
|---|---|
| Scientific title | National snakebite project on capacity building of health system on prevention and management of snakebite envenoming including its complications in selected districts of Maharashtra and Odisha in India |
| Countries of recruitment | India |
| Health condition(s) or problem(s) studied | Snakebite Envenoming |
| Intervention(s) | Community Intervention: Educational talks, IEC activities for increasing knowledge about snakes and snakebites and awareness regarding methods of preventing snakebites and early referral to health facilities. Engagement of local self-government institutions, teachers, Anganwadi, forest officials for promoting snakebite envenoming awareness among the community<br>Health System Intervention: Periodic training of Medical Officers and other healthcare workers regarding implementation of the Standard Treatment Guidelines (2017) and notification of snakebite cases |
| Key inclusion and exclusion criteria | 1. Focus Group Discussions<br>Inclusion: Member of community above 18 years of age and willing to participate<br>Exclusion: Healthcare providers<br>2. Interview and training of Medical Officers (MO) and other Healthcare workers in the block<br>All the Medical Officers and ASHA, ANM and MPW in the study blocks will be eligible<br>3. Facility Survey<br>All the Primary Health Centers, Rural Hospitals, Community Health Centers, Sub District Hospitals and District Hospitals in the selected blocks will be included. Sub centers will be excluded. |
| Study type | Type of study: Interventional<br>Study Design: Exploratory study with Retrospective, Prospective, Cross Sectional and Qualitative component conducted in community and hospital setting. |
| Date of first enrolment | Enrolment yet to begin |
| Target sample size | 1. A total of 6 FGDs (3 males, 3 females) will be conducted in each block. Each FGD will be restricted to 8 to 16 participants. Overall, 24 FGDs will be |

| Data Category | Information |
| --- | --- |
| | conducted but the number of FGDs will vary according to the saturation of responses.<br>2. Each block will have approximately 40 Medical Officers (MOs); so nearly 160 MOs will be trained during the study. Training will also be provided to outreach healthcare workers. Approximately 150 workers per block will be trained so a total of 600 workers will be trained in four blocks. Actual numbers will vary and the same MOs or healthcare workers may not be available throughout the study duration as there are frequent transfers in the health system.<br>3. FGDs will be conducted in the study areas to evaluate the post-intervention community knowledge with similar methodology as described in point no. 1 |
| Recruitment status | Pending: participants are not being recruited or enrolled at any site |
| Primary outcome(s) | Outcome Name: Case fatality rate<br>Method of measurement: Prospective data collection at public health facilities in the block followed by comparison of collected data with retrospective data of the last two years before study initiation<br>Time point: Two years from the start of the study |
| Key secondary outcomes | 1. Outcome Name: Incidence of snakebite cases<br>Method of measurement: Prospective data collection at public health facilities in the block followed by comparison of collected data with retrospective data of the last two years<br>Time point: Two years from the start of the study<br><br>2. Outcome Name: Incidence of complications due to snakebite envenoming<br>Method of measurement: Prospective data collection at public health facilities in the block followed by comparison of collected data with retrospective data of the last two years<br>Time point: Two years from the start of the study<br><br>3. Outcome Name: Implementation of the Standard Treatment Guidelines, 2017 for the management of snakebite envenoming<br>Method of measurement: Facility check surveys, periodic supervisory visits by Master trainers and subject experts<br>Time point: 12 months in the study onwards |

| Data Category | Information |
|---|---|
| Ethics Review | Ethics approval was obtained from the Institutional Ethics committee of ICMR- NIRRCH (Ref. No. D/ICEC/Sci-194/209/2021) and ICMR-RMRCB (ICMR-RMRCB/IHEC-2021/79). The study is registered under the Clinical Trial Registry India, registration number - CTRI/2021/11/038137.<br><br>Ethics committee contact –<br>Dr. Beena Joshi<br>NIRRCH Ethics Committee for Clinical Studies<br>National Institute for Research in Reproductive and Child Health<br>J.M.Street, Parel. Mumbai 400 012<br>Tel.No.: Direct- 022-24192043 / 022-24192147<br>Board no.- 022-24192000<br> |
| Completion date | NA |
| Summary Results | NA |
| IPD sharing statement | The deidentified individual clinical trial participant-level data will not be shared. PI will have ultimate authority over datasets and access. |

Data Set Source – World Health Organization Trial Registration Data Set (Version 1.3.1). https://www.who.int/clinical-trials-registry-platform/network/who-data-set. Accessed February 02, 2022.
