## Supplementary material for "National snakebite project on capacity building of health system on prevention and management of snakebite envenoming including its complications in selected districts of Maharashtra and Odisha in India: a study protocol": S7 SPIRIT_Figure

| STUDY PERIOD (months) | | | | | | | | | | |
|---|---|---|---|---|---|---|---|---|---|---|
| TIMEPOINT | -24 | 0 | 3 | 6 | 9 | 12 | 15 | 18 | 21 | 24 |
| Activities | | | | | | | | | | |
| Retrospective data | ◆— | —◆ | | | | | | | | |
| Pre-Intervention Focus Group Discussions | | | | ✓ | | | | | | |
| Facility Check Survey | | | ✓ | | | | | | | |
| Interviews of Medical Officers | | | | ✓ | | | | | | |
| Prospective data | | | | | | ◆— | — | — | — | —◆ |
| Post-Intervention Focus Group Discussions | | | | | | | | ✓ | | |
| Interventions | | | | | | | | | | |
| IEC Campaign | | | | ◆— | — | — | —◆ | | | |
| Community Talks | | | | | ◆— | — | —◆ | | | |
| Training of trainers | | | | | ◆—◆ | | | | | |
| Healthcare Provider Training | | | | | | ◆— | —◆ | | | |
| Periodic Visits to Health Facilities | | | | | | | ✓ | ✓ | ✓ | ✓ |
| Impact Assessment | | | | | | | | | | |
| Case Fatality Rate | | | | | | | | | | ✓ |
| Snakebite incidence | | | | | | | | | | ✓ |
| Complications due to SBE | | | | | | | | | | ✓ |
| Notification of snakebite cases | | | | | | | | | | ✓ |
| Implementation of Standard Treatment Guideliens, 2017 | | | | | | | ◆— | — | — | —◆ |
